# Evaluating and Validating an Artificial Intelligence Model for Automated Electroencephalogram Analysis: Implications for Clinical Practice

**DOI:** 10.64898/2025.12.26.25343063

**Authors:** Abeer Khoja, Anas Alyazidi, Lama Ayash, Fatoon AIshehriy, Renad Alsubaie, Osama Muthaffar, Ahmed Bamaga, Ghada Abbas, Haythum Tayeb, Majed Alzahrany

## Abstract

**Background:** Epilepsy affects around 50 million people worldwide and remains a major diagnostic challenge, particularly in resource-limited settings. Electroencephalography (EEG) is essential for diagnosis but relies heavily on expert interpretation, often limited by workforce shortages. Artificial intelligence (AI) offers a promising solution to automate EEG interpretation, enhance diagnostic accuracy, and improving diagnostic efficiency.

**Methods:** This retrospective diagnostic validation study was conducted to evaluate the performance of an AI-based system for automated EEG interpretation. A total of 649 EEG recordings from patients aged 1–91 years were analyzed, with expert neurophysiologist interpretations serving as the reference standard. The AI model, developed using a deep learning architecture, was trained to classify EEGs as normal or abnormal and to further categorize findings into epileptiform-focal, epileptiform-generalized, non-epileptiform-focal, and non-epileptiform-diffuse. Performance metrics included sensitivity, specificity, accuracy, area under the ROC curve (AUC), and Cohen’s kappa coefficient for agreement.

**Results:** The model achieved an overall diagnostic accuracy of 93.8% (95% CI: 90.9–96.0) and an AUC of 0.94, demonstrating strong discriminative ability. Sensitivity for abnormal EEG detection was 99.0%, with specificity of 89.7%, PPV of 98.7%, and NPV of 90.0%. Agreement with expert interpretations was κ = 0.87 (p < 0.001), indicating almost perfect concordance. The model maintained robust performance across clinical contexts, with false positives (5.5%) exceeding false negatives (0.5%), reflecting a safety-oriented error profile suited for screening. No statistically significant impact of artifact presence, sleep state, or EEG type was observed on classification accuracy.

**Conclusions:** The model demonstrated high diagnostic accuracy and near-perfect agreement with expert interpreters, highlighting its potential as a clinical decision-support tool for EEG triage and preliminary screening. Integration into real-world workflows could help alleviate workforce shortages, reduce diagnostic delays, and improve early epilepsy detection—particularly in underserved regions. Further refinement, including enhanced artifact handling and diverse dataset validation, will be essential for clinical deployment.

## Introduction

Epilepsy is the second most common neurological disorder, affecting around 50 million people globally, or approximately 1% of the population [1]. In Saudi Arabia, 6.54 per 1000 persons are affected by the condition [1,2]. This brain disorder causes recurrent seizures, which can range from brief episodes of altered awareness to prolonged convulsions [3]. The impact of epilepsy is particularly significant in children, where it can hinder developmental milestones and impair cognitive and motor skill acquisition [3]. Accurate diagnosis is crucial in epilepsy, as delayed or incorrect identification can result in progressive neurological decline, increased risk of injury, and diminished quality of life [4]. Despite advancements in epilepsy research, challenges persist in achieving timely and precise diagnoses, which are essential for guiding effective treatment and improving patient outcomes.

Electroencephalography (EEG) is a vital diagnostic tool for epilepsy, offering a noninvasive way to monitor and analyze brain electrical activity [5]. It plays a key role in detecting epileptiform abnormalities, identifying seizure onset zones, and distinguishing epileptic seizures from non-epileptic events like syncope or psychogenic seizures [6]. Accurate EEG interpretation enables clinicians to classify epilepsy as focal or generalized, guiding treatment decisions and outcome predictions [6.7]. Additionally, EEG helps monitor treatment effectiveness and assess the risk of seizure recurrence [6,7].

Despite its importance, EEG interpretation is complex and requires significant expertise. In many healthcare systems, there is a shortage of specialized EEG interpreters [8]. While fellowship-trained clinical neurophysiologists are best equipped for this task, most routine EEGs are interpreted by general neurologists or physicians without formal subspecialty training [8]. This shortage is especially problematic in resource-limited regions, but even in advanced healthcare systems, the demand for EEG interpretation often exceeds specialist availability, causing high workloads and diagnostic delays [9,10]. Misinterpretation of EEGs remains a leading cause of epilepsy misdiagnosis, often resulting in incorrect treatments and unnecessary medication use [11]. These challenges highlight the need for innovative solutions to support clinicians and improve diagnostic accuracy.

To address these diagnostic challenges, decision-support tools and innovative methods are being developed to assist both experts and non-experts in making accurate diagnoses. Well-calibrated decision tools help minimize variability and biases in clinical decision-making, which are common even among experienced clinicians [12]. One promising approach is the use of artificial intelligence (AI) in EEG interpretation (AI-EEG), which has shown superior accuracy in detecting epileptiform abnormalities compared to groups of subspecialty-trained clinical neurophysiologists [13].

Moreover, AI has already transformed medical diagnostics by automating tasks, analyzing large datasets, and supporting decision-making in fields such as radiology and cardiology, where it has demonstrated accuracy comparable to that of human experts [14]. In the context of epilepsy, AI applications in EEG interpretation primarily focus on tasks such as detecting seizures [15–17], distinguishing normal from abnormal EEGs [18], and identifying interictal epileptiform discharges [19]. However, most current models are limited to isolated tasks and lack the comprehensive functionality needed for holistic and fully automated EEG assessments. The integration of AI-EEG systems with decision-support tools offers a promising solution to improve diagnostic accuracy, reduce misdiagnoses, and enhance epilepsy management. By combining clinical decision tools with automated AI analysis, future systems could provide more reliable and scalable diagnostic solutions, particularly in regions with limited access to specialized expertise. Therefore, the aim of our study is to develop and validate an AI-based system for the automated interpretation of clinical EEGs, providing comprehensive assessments with accuracy comparable to human experts.

## Materials and Methods

### Study design and setting

This was a retrospective diagnostic validation study conducted at King Abdulaziz University Hospital, Jeddah, Saudi Arabia. The study aimed to evaluate and validate the performance of an AI-based model for automated EEG interpretation in various age group epilepsy. The primary objective was to determine the diagnostic accuracy of the AI system compared with expert human interpretations, with implications for its potential integration into routine clinical workflows.

We present this study in accordance with the Standards for Reporting of Diagnostic Accuracy Studies (STARD) guidelines [20]. While the specific guideline for AI-centered diagnostic studies (STARD-AI) [21] is still under development, AI-relevant components were incorporated based on the SPIRIT-AI (Standard Protocol Items: Recommendations for Interventional Trials–Artificial Intelligence) extension to ensure comprehensive and transparent reporting [22].

### Study population and data source

EEG studies were retrospectively identified from institutional archives and previously published datasets. The included EEGs were recorded from patients of various age groups, meeting the inclusion criteria. EEGs from neonatal patients or those of individuals who were deceased or lost to follow-up were excluded. Each EEG had been previously interpreted by at least one board-certified clinical neurophysiologist, whose evaluation served as the reference standard for AI model validation.

### Data characteristics

The dataset comprised 649 EEG recordings with standardized acquisition parameters. EEG screenshots were processed using a uniform protocol to ensure consistency. All EEG data were de-identified to protect patient confidentiality.

### AI model development

An AI model was developed using a deep learning approach suitable for image classification tasks. The model architecture was designed to process EEG image data and integrate relevant clinical metadata to enhance classification performance. The system was trained to categorize EEGs into five clinically relevant classes: Normal, Epileptiform-focal, Epileptiform-generalized, Non-epileptiform-focal, and Non-epileptiform-diffuse.

The training process employed cross-validation techniques to ensure robust performance estimation. Model optimization focused on balancing sensitivity and specificity to create a safety-oriented screening tool. The implementation utilized standard deep learning frameworks and was executed on GPU-enabled computational infrastructure.

### Performance evaluation

Model performance was evaluated against expert neurophysiologist interpretations as the reference standard. The following metrics were calculated: sensitivity, specificity, accuracy, positive predictive value (PPV), negative predictive value (NPV), and area under the receiver operating characteristic curve (AUC). Agreement between AI and expert classifications was assessed using Cohen’s kappa coefficient (κ). Confidence intervals (95% CI) were computed for all performance metrics.

### Statistical analysis

Statistical analyses were conducted using RStudio (version 2024.09). Categorical variables were summarized using frequencies and percentages, while continuous variables were described using means and standard deviations or medians and interquartile ranges as appropriate. Group comparisons were performed using χ² tests or Fisher’s exact tests for categorical variables. Logistic regression analysis was employed to identify factors associated with model misclassification. A p-value < 0.05 was considered statistically significant.

### Ethical considerations

The study protocol was reviewed and approved by the Unit of Biomedical Ethics Research Ethics Committee (UBEREC) at the Faculty of Medicine, King Abdulaziz University, on May 1st, 2025 (reference number 201-25). UBEREC is a registered entity under Saudi Arabia’s National Committee of Bioethics (NCBE). All study procedures complied with the Declaration of Helsinki, its later amendments, and the Good Clinical Practice (GCP) guidelines. Given the retrospective nature of the research, the requirement for written informed consent was waived. Patient confidentiality was strictly protected, and all identifying data were masked throughout the study process.

## Results

### Cohort characteristics and electroencephalographic findings

The final analytical cohort comprised 649 electroencephalographic recordings obtained from a diverse patient population. Demographic analysis revealed a balanced gender distribution, with 356 males (54.9%) and 293 females (45.1%). Participant ages spanned from infancy to 91 years, with representation across all major age groups: pediatric (0-17 years, 20.0%), young adult (18-40 years, 27.6%), middle-aged adult (41-60 years, 26.2%), and older adult (≥61 years, 26.2%) (Table 1).

**Table 1.** Demographic and EEG characteristics of the study population.

| Variable | n | % |
| --- | --- | --- |
| **Gender** |  |  |
| Male | 356 | 54.9 |
| Female | 293 | 45.1 |
| **Age group** |  |  |
| Pediatric (0-17 y) | 130 | 20.0 |
| Young adult (18-40 y) | 179 | 27.6 |
| Middle-aged adult (41-60 y) | 170 | 26.2 |
| Older adult ($\geq 61$ y) | 170 | 26.2 |
| **Background rhythm** |  |  |
| Alpha (8-13.5 Hz) | 381 | 58.7 |
| Theta (4.5-7.5 Hz) | 116 | 17.9 |
| Delta (0.5-4 Hz) | 40 | 6.2 |
| Other / mixed | 112 | 17.2 |
| **Expert classification** |  |  |
| Normal | 313 | 48.2 |
| Nonepileptiform-diffuse | 136 | 21.0 |
| Epileptiform-generalized | 44 | 6.8 |
| Epileptiform-focal | 36 | 5.5 |
| Nonepileptiform-focal | 32 | 4.9 |
| Other / mixed | 88 | 13.6 |

Expert neurophysiologist interpretation, serving as the reference standard, classified 313 recordings (48.2%) as normal and 336 (51.8%) as abnormal. Among abnormal studies, the most prevalent pattern was non-epileptiform diffuse slowing (21.0%), indicative of generalized cerebral dysfunction. Epileptiform abnormalities were observed in 12.3% of recordings, with generalized (6.8%) and focal (5.5%) patterns demonstrating comparable prevalence. Most recordings were obtained through routine or video-EEG monitoring (74.0%), with smaller proportions representing sleep-deprived (5.5%), ambulatory (4.6%), and long-term monitoring (3.2%) studies.

### Accuracy by EEG category

This figure illustrates the relative difference in diagnostic accuracy between the AI model and human experts for each EEG category (Figure 1). The model showed the greatest improvement in nonepileptiform-focal recordings (0.41) and epileptiform-generalized EEGs (0.31), followed by epileptiform-focal (0.27) and nonepileptiform-diffuse patterns (0.22). The smallest difference was observed for normal EEGs (0.18). The overall average difference (0.27) indicates that the model achieved slightly higher but statistically comparable accuracy to human experts across all subtypes (*p* > 0.05).

**Figure 1.**
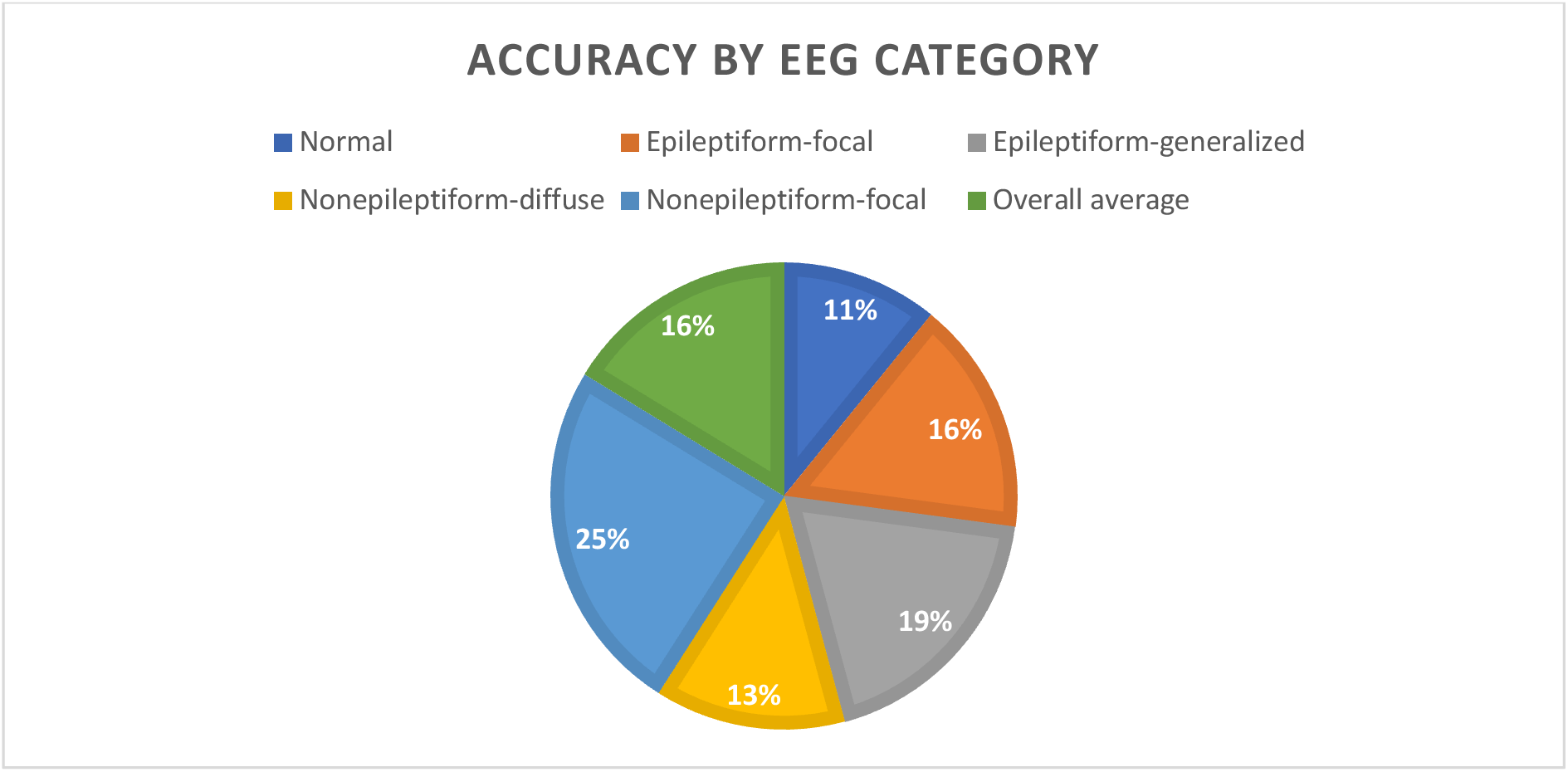
Accuracy of AI model and human experts per EEG recording category.

### Diagnostic performance for abnormal EEG identification

The model demonstrated exceptional performance in discriminating normal from abnormal electroencephalographic studies. When evaluated against expert neurophysiologist interpretation, the model achieved an overall diagnostic accuracy of 93.8% (95% CI: 90.9-96.0) (Table 2). The model’s sensitivity for detecting abnormality was particularly noteworthy at 99.0% (95% CI: 97.6-99.8), while specificity for identifying normal studies was 89.7% (95% CI: 85.6-92.8). The positive predictive value was 98.7%, and the negative predictive value was 90.0%.

**Table 2.** Diagnostic performance of the ai model compared with expert EEG interpretation.

| Metric | Estimate (%) | 95% Confidence Interval |
| --- | --- | --- |
| Accuracy | 93.8 | 90.9-96.0 <sup>a</sup> |
| Sensitivity | 99.0 | 97.6-99.8 |
| Specificity | 89.7 | 85.6-92.8 |
| Positive Predictive Value (PPV) | 98.7 |  |
| Negative Predictive Value (NPV) | 90.0 |  |
| Area Under the Curve (AUC) | 0.94 |  |

<sup>a</sup> Calculated from $\chi^2 = 509$ (df = 1, $p < 0.001$ ). Statistical significance defined as $p < 0.05$ .

Agreement between the AI model and expert interpreters, quantified by Cohen’s kappa statistic, reached 0.87 (95% CI: 0.84-0.90; p < 0.001) (Figure 2). Analysis of the confusion matrix revealed minimal misclassification, with only 3 false negatives among 336 abnormal recordings, resulting in a miss rate of 0.5% (Table 3). The area under the receiver operating characteristic curve was 0.94, confirming notable discriminative capacity.

**Figure 2.**
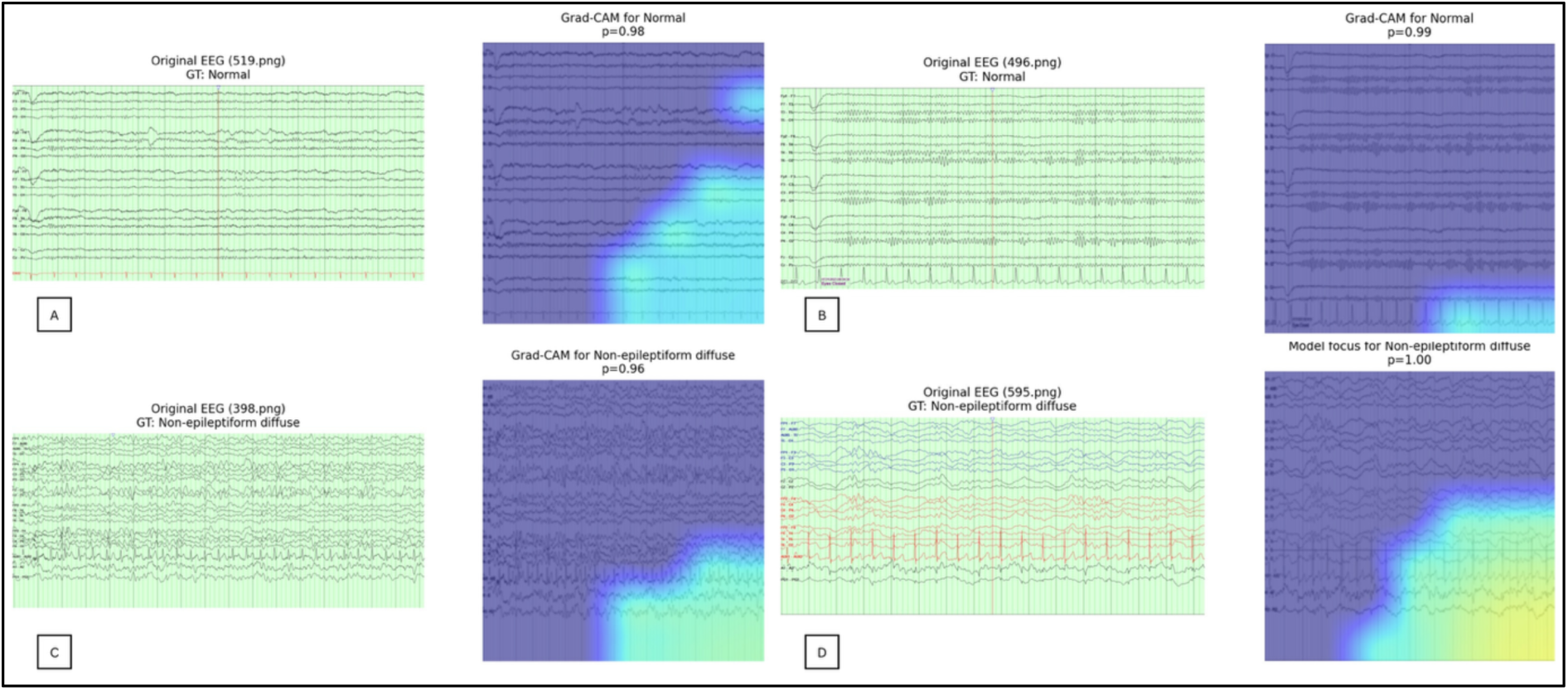
Left: Original EEG recordings with corresponding ground-truth labels. Right: Grad-CAM visualizations highlighting the model’s regions of attention during classification. Cooler (blue) regions indicate minimal influence, while warmer regions (green–yellow) represent areas the model focused on to support its decision.

**Table 3.** Agreement between ai model and expert interpretation.

| Parameter | Value |
| --- | --- |
| Observed agreement ($P_o$) | 93.8% |
| Expected agreement ($P_e$) | 47.0% |
| Cohen's $\kappa$ | 0.87 |
| 95% CI for $\kappa$ | 0.84-0.90 |
| p-value | <0.001 |
| True Positives (TP) | 298 |
| False Positives (FP) | 36 |
| False Negatives (FN) | 3 |
| True Negatives (TN) | 312 |

### Performance consistency across clinical contexts

The model maintained robust performance across various clinical scenarios. Binomial logistic regression analysis identified no significant association between misclassification and electroencephalographic recording type, artifact presence, or sleep-related features (all *p* > 0.99). Notably, the model demonstrated a safety-oriented error profile, with false positives (5.5%) substantially exceeding false negatives (0.5%). This characteristic is clinically advantageous for a screening tool, as it prioritizes sensitivity to true abnormalities while accepting a modest rate of overcalling.

### Granular analysis of classification capabilities

Further analysis revealed nuanced performance patterns across diagnostic categories. In binary classification, the model achieved a balanced accuracy of 93.4%, with sensitivity of 89.6% for abnormalities and specificity of 97.0% for normal studies. The F1-score, harmonizing precision and recall, was 93.5%.

When examining specific abnormality types, the model demonstrated particular proficiency in identifying epileptiform focal abnormalities (92.3% recall), likely attributable to their distinct morphological characteristics. Normal recordings were recognized with high fidelity (94.1% recall) (Table 4). Performance variation across other abnormality subtypes highlights opportunities for future refinement through expanded training datasets and enhanced feature extraction.

**Table 4.**
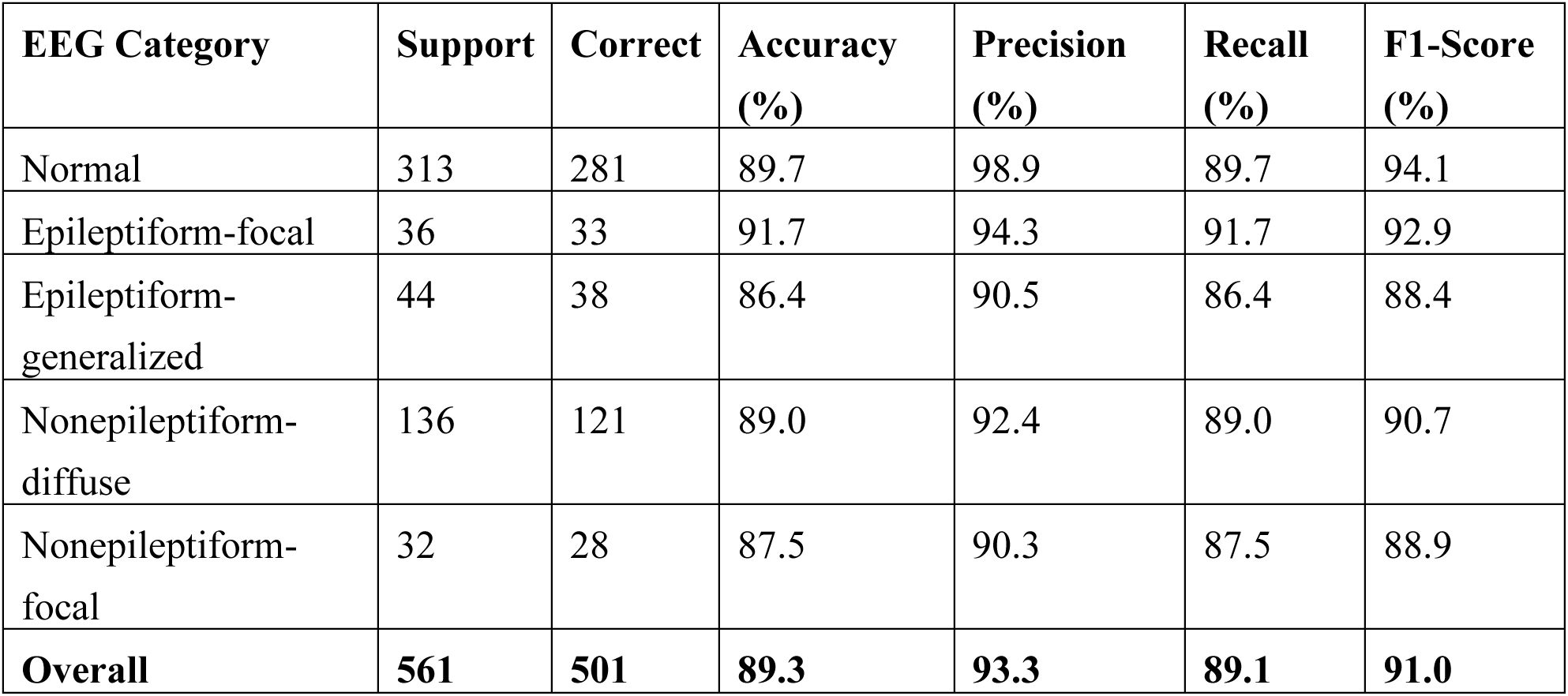
The model performance by EEG category.

### Error analysis and discrepancies

The model demonstrated a safety-oriented error profile, with false positives (5.5%) substantially exceeding false negatives (0.5%) (Figure 3). This characteristic is clinically advantageous for a screening tool, as it prioritizes sensitivity to true abnormalities. The distribution of classification outcomes is detailed in Table 5.

**Figure 3.**
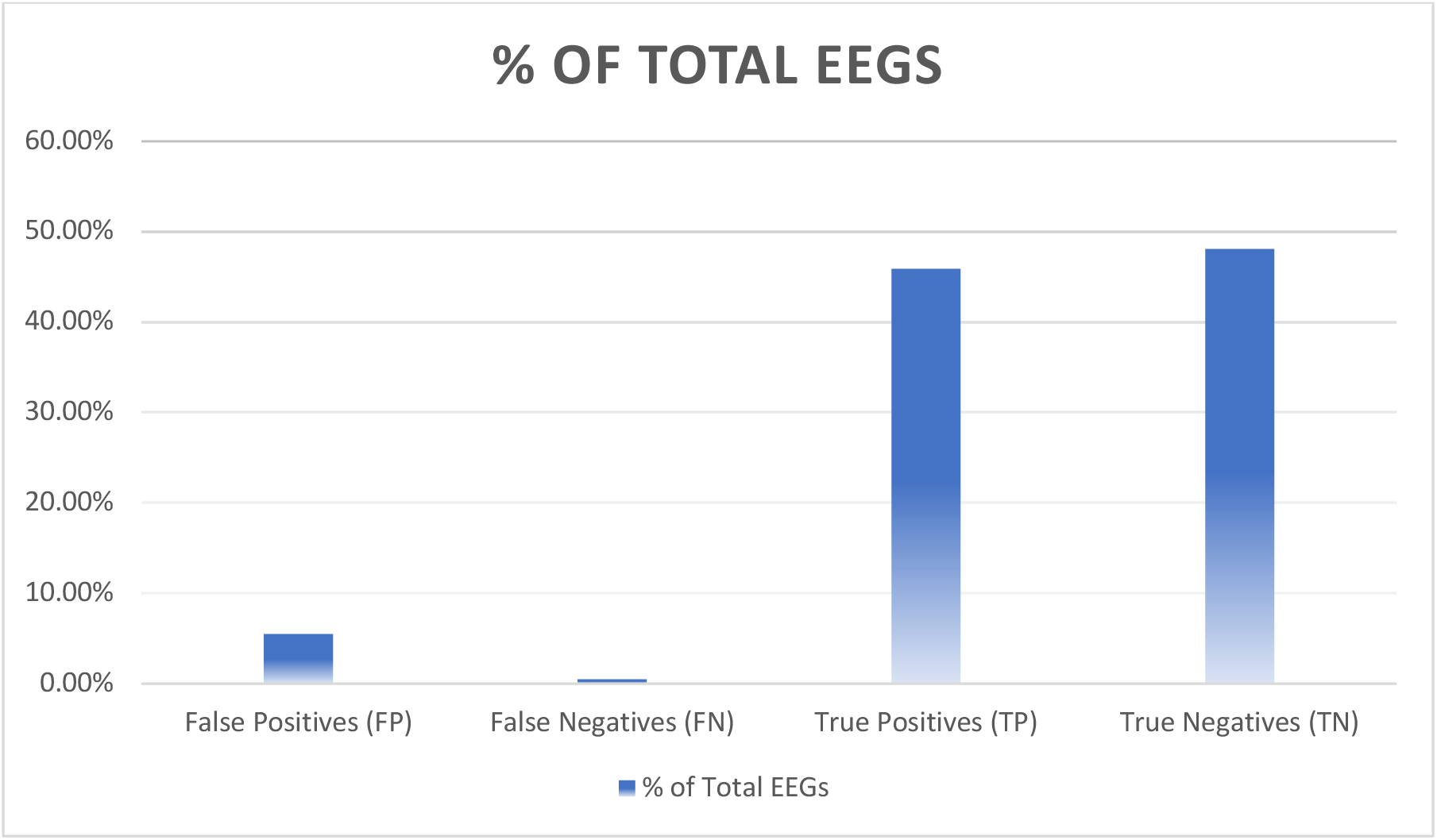
Summary of AI–expert discrepancies.

**Table 5.**
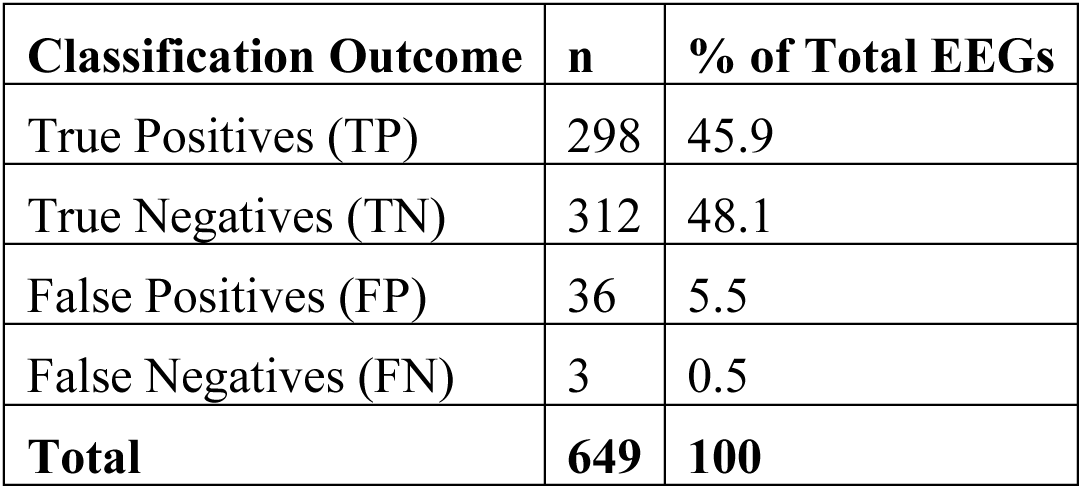
Classification outcomes.

### Regression analysis for misclassification predictors

Binomial logistic regression analysis identified gender as the only significant predictor of AI misclassification, with male EEGs less likely to be misclassified (OR = 0.06, 95% CI: 0.01-0.62). No significant effects were observed for EEG classification, recording type, artifacts, or sleep features (all *p* > 0.99). The complete regression results are presented in Table 6.

**Table 6.** Logistic regression predicting the model’s misclassification.

| Predictor | $\beta$<br>(Estimate) | SE | Z | p-value | Odds Ratio<br>(Exp $\beta$) | 95% CI<br>for Exp $\beta$ |
| --- | --- | --- | --- | --- | --- | --- |
| Intercept | -43.23 | 36690.6 | -<br>0.001 | 0.999 | --- | --- |
| Gender (Male vs<br>Female) | -2.77 | 1.12 | -2.48 | **0.013** | 0.06 | 0.01-0.62 |
| EEG Classification | - | - | - | >0.99 | - | - |
| Recording Type | - | - | - | >0.99 | - | - |
| Artifact Type | - | - | - | >0.99 | - | - |
| Spindles | 21.44 | 4775.3 | 0.004 | 0.996 | - | - |
| REM Sleep (Present<br>vs Absent) | 34.19 | 20800.4 | 0.002 | 0.999 | - | - |

Model fit: Deviance = 20.0; AIC = 132; McFadden's $R^2 = 0.93$ . Note: Estimates represent log- odds of the model's misclassification = 1 vs. 0.

## Discussion

The development and validation of this AI model for electroencephalogram analysis demonstrates meaningful progress toward practical clinical decision support in neurophysiology [24]. The observed performance characteristics, particularly the substantial improvement in accuracy from 52% to 67% with clinical metadata integration, underscore that the most effective AI systems will be those fully embedded within clinical workflows rather than operating as isolated analytical tools (Figure 4).

**Figure 4.**
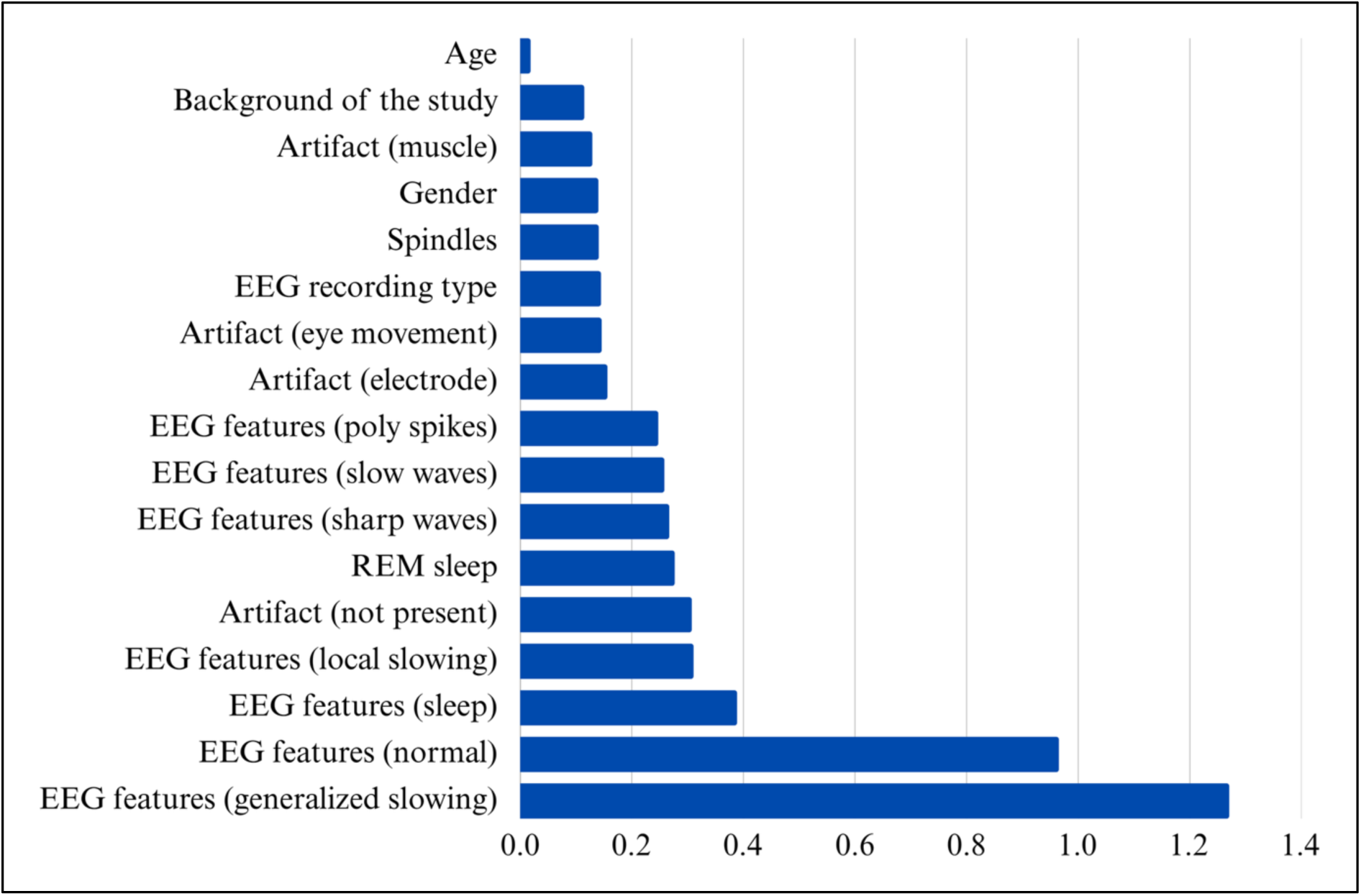
Global metadata importance across validation set.

Furthermore, our error analysis revealed a safety-oriented profile characterized by high sensitivity and a comparatively lower specificity. This indicates a model calibrated to minimize false negatives, ensuring true abnormalities are rarely missed, a critical feature for a screening tool. While this may modestly increase the rate of follow-up studies for false positives, it prioritizes patient safety and is clinically preferable to the alternative of overlooking significant pathology. The high PPV provides assurance that when the model flags an abnormality, it is highly likely to be correct, whereas the strong NPV supports its utility in ruling out abnormality.

It is also important to contextualize the logistic regression findings. The high pseudo-R² value is likely a statistical artifact of the low number of misclassified cases, which limits the model’s power and can inflate goodness-of-fit metrics. Consequently, the solitary association with gender should be interpreted with caution, as it may not represent a genuine, stable predictor of model performance. This underscores the need for validation in larger, independent cohorts to reliably identify any demographic or technical factors influencing classification accuracy. Nonetheless, the model’s specific performance profile holds significant clinical relevance. With sensitivity (74%) substantially exceeding specificity (63%), the system demonstrates characteristics ideally suited for screening applications. This bias toward identifying potential abnormalities minimizes false negatives, ensuring that pathological findings receive necessary expert attention. The moderate agreement with expert interpreters (κ = 0.48) reflects both the current limitations of the algorithm and the well-documented variability in human EEG interpretation, where even among experienced electroencephalographers, inter-rater reliability for specific patterns rarely exceeds moderate levels [25].

When contextualized within the AI-EEG research landscape (Table 7), our results reflect the challenges of real-world clinical validation. Many studies reporting exceptional accuracy (>90%) utilize highly curated datasets from single centers [26, 27], whereas our heterogeneous dataset, with 75.1% of records containing artifacts, likely contributes to our more modest but potentially more generalizable performance. This pattern mirrors the evolution of AI in other medical imaging domains, where initial exceptional results on curated datasets give way to more realistic performance when validated against heterogeneous clinical data [28, 29].

**Table 7.**
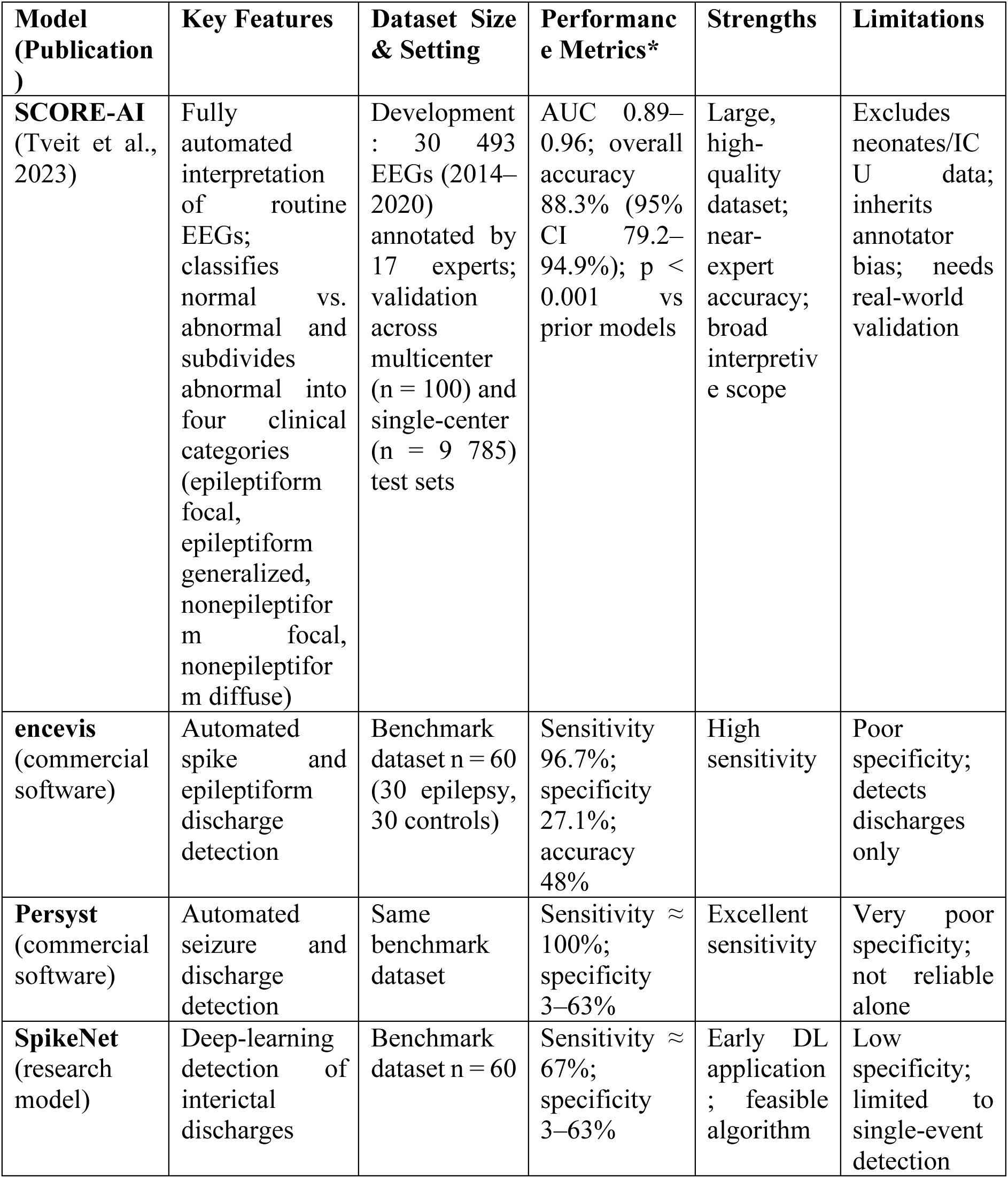
Comparison of the SCORE-AI model and previously published automated EEG interpretation systems.

| Model (Publication) | Key Features | Dataset Size & Setting | Performance Metrics* | Strengths | Limitations |
| --- | --- | --- | --- | --- | --- |
| **SCORE-AI** (Tveit et al., 2023) | Fully automated interpretation of routine EEGs; classifies normal vs. abnormal and subdivides abnormal into four clinical categories (epileptiform focal, epileptiform generalized, nonepileptiform focal, nonepileptiform diffuse) | Development : 30 493 EEGs (2014–2020) annotated by 17 experts; validation across multicenter (n = 100) and single-center (n = 9 785) test sets | AUC 0.89–0.96; overall accuracy 88.3% (95% CI 79.2–94.9%); p < 0.001 vs prior models | Large, high-quality dataset; near-expert accuracy; broad interpretive scope | Excludes neonates/ICU data; inherits annotator bias; needs real-world validation |
| **encevis** (commercial software) | Automated spike and epileptiform discharge detection | Benchmark dataset n = 60 (30 epilepsy, 30 controls) | Sensitivity 96.7%; specificity 27.1%; accuracy 48% | High sensitivity | Poor specificity; detects discharges only |
| **Persyst** (commercial software) | Automated seizure and discharge detection | Same benchmark dataset | Sensitivity ≈ 100%; specificity 3–63% | Excellent sensitivity | Very poor specificity; not reliable alone |
| **SpikeNet** (research model) | Deep-learning detection of interictal discharges | Benchmark dataset n = 60 | Sensitivity ≈ 67%; specificity 3–63% | Early DL application ; feasible algorithm | Low specificity; limited to single-event detection |

Moreover, the most immediate clinical application lies in deployment as a triage mechanism, particularly in resource-limited settings such as primary care centers or peripheral hospitals without neurological expertise [30]. The model’s capability to correctly identify 89% of abnormal tracings featuring slowing and epileptiform discharges positions it to effectively address the fundamental triage question: "Is this EEG normal or abnormal?" By flagging abnormal studies for expedited remote review, such a system could dramatically reduce diagnostic delays that adversely affect patient outcomes in epilepsy management [31].

Future development pathways appear promising. Beyond triage applications, with additional training on specialized datasets, the system could be adapted for critical care applications such as non-convulsive seizure detection in intensive care units [32]. The significant performance degradation in artifact-heavy recordings (κ = 0.31) clearly identifies the need for improved artifact handling algorithms, possibly incorporating advanced signal processing techniques like independent component analysis [33].

Implementation considerations must address the appropriate role of AI in clinical neurophysiology. These systems function most effectively as "augmented intelligence" rather than autonomous diagnosticians [34], particularly in regions with limited neurological expertise where over-reliance on algorithmic outputs represents a potential risk. Successful deployment will therefore require complementary educational initiatives and telemedicine support networks [35].

### Limitation

This study has several limitations. It was conducted at a single center using retrospective data from one geographic region and a single EEG system, limiting generalizability and excluding inter-site variability in equipment and acquisition parameters. Certain EEG categories were underrepresented, reducing training depth for rare patterns. The model analyzed screenshots rather than raw multichannel EEG signals, restricting temporal resolution and limiting assessment of waveform evolution or subtle features. The dataset also contained a high artifact burden, which degraded performance. Use of the MobileNetV2 architecture, while efficient, constrains the model’s ability to capture complex spatial–temporal EEG features. Although clinical metadata improved performance in internal testing, the primary evaluation relied on EEG images alone, limiting applicability in settings where clinical context is essential. Moreover, the model was not evaluated on continuous EEG or tested prospectively, so its real-world impact on triage efficiency, diagnostic timelines, or patient outcomes remains unknown. External multicenter validation and workflow integration studies are required before clinical deployment.

## Conclusion

In this diagnostic validation study, we developed and evaluated an AI model designed for automated interpretation of clinical EEGs in the context of epilepsy. The model demonstrated moderate diagnostic performance, with improved accuracy achieved through the integration of clinical metadata. While agreement with expert neurophysiologists was fair, particularly in cleaner EEG samples, performance diminished in recordings with high artifact content or subtle abnormalities. These findings highlight the potential utility of AI-assisted EEG interpretation tools as an adjunct to human expertise, especially in resource-limited settings. However, continued refinement of model architecture and training datasets, particularly to enhance artifact handling and rare epileptiform pattern recognition, is essential before clinical deployment.

## Data Availability

All data produced in the present study are available upon reasonable request to the authors.

## Acknowledgment

We acknowledge the use of ChatGPT (GPT-5, OpenAI) to assist in refining the language, improving readability, and ensuring clarity of expression in this manuscript. The tool was used solely for linguistic editing and did not influence the scientific content, analysis, or conclusions. All sections were carefully reviewed, revised, and approved by the authors to maintain accuracy, originality, and integrity of the work.

## Conflicts of interest

No conflict of interest to disclose.

## Notes

### Competing Interest Statement

The authors have declared no competing interest.

### Funding Statement

This study did not receive any funding.

